# Distress symptoms of old age and mild cognitive impairment are two distinct dimensions in older adults without major depression: effects of adverse childhood experiences and negative life events

**DOI:** 10.1101/2023.11.03.23297890

**Authors:** Vinh-Long Tran-Chi, Michael Maes, Gallayaporn Nantachai, Solaphat Hemrungrojn, Marco Solmi, Chavit Tunvirachaisakul

**Affiliations:** Ph.D. Program in Clinical Sciences, School of Global Health, Faculty of Medicine, Chulalongkorn University, Bangkok, Thailand; Department of Psychiatry, Faculty of Medicine, Chulalongkorn University, Bangkok, Thailand; Research Institute, Medical University of Plovdiv, Plovdiv, Bulgaria; Department of Psychiatry, Medical University of Plovdiv, Plovdiv, Bulgaria; Sichuan Provincial Center for Mental Health, Sichuan Provincial People’s Hospital, School of Medicine, University of Electronic Science and Technology of China, Chengdu, China; Key Laboratory of Psychosomatic Medicine, Chinese Academy of Medical Sciences, Chengdu, China; Kyung Hee University, 26 Kyungheedae-ro, Dongdaemun-gu, Seoul, South Korea; Cognitive Impairment and Dementia Research Unit, Department of Psychiatry, Faculty of Medicine, Chulalongkorn University, Bangkok, Thailand; Cognitive Fitness and Biopsychiatry Technology Research Unit, Faculty of Medicine, Chulalongkorn University, Bangkok, Thailand; Somdet Phra Sungharaj Nyanasumvara Geriatric Hospital, Department of Medical Services, Ministry of Public Health, Chon Buri Province, Thailand; Department of Psychiatry, University of Ottawa, Ontario, Canada; Regional Centre for the Treatment of Eating Disorders and On Track, The Champlain First Episode Psychosis Program, Department of Mental Health, The Ottawa Hospital, Ontario, Canada; Ottawa Hospital Research Institute (OHRI), Clinical Epidemiology Program, University of Ottawa, Ottawa, Ontario, Canada; Department of Child and Adolescent Psychiatry, Charité Universitätsmedizin, Berlin, Germany

**Keywords:** depression, adverse childhood experiences, mild cognitive impairments, affective disorders, negative life events, neurocognitive deficits

## Abstract

**Background:** Studies in old adults showed bidirectional interconnections between amnestic mild cognitive impairment (aMCI) and affective symptoms and that adverse childhood experiences (ACE) may affect both factors. Nevertheless, these associations may be confined to older adults with clinical depression.

**Aims:** To delineate the relationship between clinical symptoms of aMCI and affective symptoms in older adults without major depression (MDD) or dysfunctions in activities of daily living (ADL).

**Methods:** This case-control study recruited 61 participants with aMCI (diagnosed using Petersen’s criteria) and 59 older adults without aMCI and excluded subjects with MDD and ADL dysfunctions.

**Results:** We uncovered 2 distinct dimensions, namely distress symptoms of old age (DSOA) comprising anxiety, depression, perceived stress and neuroticism scores, and mild cognitive dysfunctions (mCoDy) comprising episodic memory test scores, and the total Mini-Mental State Examination (MMSE) and Montreal Cognitive Assessment (MoCA) scores. A large part of the variance (37.9%) in DSOA scores was explained by ACE, negative life events (health and financial problems), a subjective feeling of cognitive decline, and education (all positively). While ACE and NLE have a highly significant impact on the DSOA, they are not associated with the mCoDy scores. Cluster analysis showed that the diagnosis of aMCI is overinclusive because some subjects with DSOA symptoms may be incorrectly classified as aMCI.

**Conclusions:** The clinical impact is that clinicians should carefully screen older adults for DSOA after excluding MDD. DSOA might be misinterpreted as aMCI.

## Introduction

Mild cognitive impairment (MCI) has been observed to have a significant prevalence among the elderly population affecting approximately 10-15% of individuals aged 65 and above ^1^. Mild deficits in episodic memory, executive functions, visuospatial skills, processing speed, cognitive flexibility, and problem-solving capacity are prominent characteristics of amnestic mild cognitive impairment (aMCI), while fundamental activities associated with daily living (ADL) remain unimpaired ^2–4^. aMCI can be seen as a cognitive phase that lies between the normal aging process and the onset of dementia ^1^. It is worth noting that the annual conversion rate of aMCI to Alzheimer’s disease (AD) is approximately 16.5%, although a small proportion of aMCI patients (8%) experience a recovery from this condition ^5^.

The Consortium to Establish a Registry for Alzheimer’s Disease (CERAD) developed a neuropsychological test battery to evaluate patients with neurocognitive deficits including MCI ^6^. The CERAD tests indicate that individuals with MCI have deficits in verbal fluency tasks (VFT), as well as word list memory (WLM) and world list recognition (WLR) ^7^. Age is a crucial determinant in comprehending cognitive decline due to the frequent association between MCI and the process of aging ^7^. Moreover, education level is frequently regarded as a potential confounding variable in cognitive decline research because it can affect cognitive performance; higher levels of education are associated with greater cognitive reserve, which may delay the onset of cognitive impairment or dementia ^7,8^.

Previous research has indicated that adverse childhood experiences (ACE), encompassing various forms of abuse (physical, emotional, and sexual), neglect (physical and emotional), and family dysfunctions, are significantly correlated with later cognitive impairments ^9–11^. Negative Life Events (NLE), such as severe illness, and financial problems, may be associated with increasing cognitive decline in older adults ^12–14^.

Late-life depression is a strong risk factor for normal subjects progressing to MCI ^15^ and dementia ^14^. Several systematic reviews and meta-analyses of environmental risk factors for late-life depression have been published ^16–18^. MCI patients have higher rates of depression than normal adults, and those rates are independent from age, race, gender, and study type ^19^. A meta-analysis showed the prevalence of depression in patients with MCI is as high as 32% ^20^. Educational attainment is associated with depressive symptoms of aging ^21^, and older adults with lower education attainment showed to have higher depressive symptoms ^22^. Recently, Jirakran, Vasupanrajit, Tunvirachaisakul and Maes ^23^ detected that in adult depression one factor could be extracted from increased depression, perceived stress, neuroticism, and anxiety scores, indicating that these domains are all manifestations of the underlying construct. However, there are no data whether in older adults one factor may be extracted from stress, anxiety, depression, and neuroticism domain scores.

ACE and NLE are associated with the onset of depressive symptoms ^24^ and with higher odds of late-life depressive symptoms ^25^. Increased levels of depressive symptoms and ACE exposures are associated with an increased risk of depressive symptoms in the elderly ^26–30^. However, no research has examined whether age, educational attainment, ACE, NLE, have cumulative effects in predicting severity of mild cognitive impairments in older adults. Moreover, there are no data indicating whether in older adults, cognitive symptoms are associated with affective symptoms or whether neurocognitive and affective symptoms are independent phenomena. Furthermore, there are no data whether ACE and NLE are independently associated with the neurocognitive deficits in older adults after considering the effects of affective symptoms.

Hence, the study aims to delineate the relationship between clinical symptoms of MCI and affective symptoms, and whether in older adults without major depression, ACE and NLE predict neurocognitive deficits independently from affective and distress symptoms. In addition, we examine whether in older adults one factor may be extracted from depressive, anxiety, neuroticism, and perceived distress symptoms, and whether cognitive deficits are part of this factor.

## Material and methods

### Participants

A cross-sectional study was conducted to compare individuals with aMCI to a group of healthy control subjects. The study sample comprised individuals of both genders, with an age range spanning from 60 to 75 years. The study recruited healthy participants from Bangkok, Thailand, whereas individuals with aMCI were recruited from the Outpatient Department of the Dementia Clinic at King Chulalongkorn Memorial Hospital in Bangkok, Thailand from May 2022 to March 2023. The clinical Petersen’s criteria were utilized to diagnose aMCI in the older adult population. ^31^ These criteria involve the identification of subjective and objective memory impairments, together with the lack of dementia and alterations in activities of daily living (ADL). Furthermore, people with amnestic mild cognitive impairment (aMCI) complied with the Petersen criteria and exhibited a modified Clinical Dementia Rating (CDR) score of 0.5. The control group had a CDR score of 0 and did not meet Petersen’s criteria ^31^. The healthy older adults were recruited from the Health Check-up Clinic, members of neighbourhoods’ senior clubs, healthy elderly carers of individuals with aMCI) who were patients at the Dementia Clinic, and senior volunteers affiliated with the Red Cross.

Participants with stroke, Parkinson’s disease, any dementia subtype, multiple sclerosis, schizophrenia, bipolar disorder, autism, delirium, metabolic disorder, malaria, HIV, chronic obstructive pulmonary disease (COPD), chronic kidney disease, cancer, substance abuse, alcoholism, inability to speak or communicate, blindness or impaired vision even with corrective lenses, hearing loss, inability to sit stably due to physical conditions such as chronic pain or low back pain, and those who had undergone cognitive training within three months prior to the study were excluded from the study. Ultimately, the entirety of the subjects was allocated to either of the two study groups, consisting of 59 individuals classified as healthy controls and 61 individuals diagnosed with aMCI.

Before taking part in the study, all volunteers, and individuals were required to submit written informed consent. The study conducted in this research adhered to ethical and privacy standards that are recognized both in Thailand and internationally. These standards are in accordance with the International Guideline for the Protection of Human Subjects, as mandated by influential documents such as the Declaration of Helsinki, the Belmont Report, the International Conference of Harmonization in Good Clinical Practice, and the CIOMS Guidelines. The present study received approval from the Institutional Review Board (IRB) of the Faculty of Medicine at Chulalongkorn University in Bangkok, Thailand (No. 0372/65).

### Clinical assessments

#### Neurocognition

We used a semi-structured interview to collect socio-demographic data comprising age, sex, relationship status, year of education, and marital status. The scales used to assess cognition were the Thai Mini-Mental State Examination (MMSE) ^32^, the Thai Montreal Cognitive Assessment (MoCA) ^33^, and three rating scales of the Thai Consortium to Establish a Registry for Alzheimer’s Disease (CERAD) Neuropsychological Assessment Battery ^7^. The MMSE is a 30Lquestion assessment of cognitive function ^34^. The Thai version of the Mini-Mental State Examination was developed in 1993 and has been extensively used in Thailand to screen cognitive impairment and dementia ^32^. The scale consists of six subtests measuring orientation, registration, attention, word recall, language, and computation. The total score ranges from 0 to 30. The MoCA was developed by Nasreddine, Phillips, Bédirian, Charbonneau, Whitehead, Collin, Cummings and Chertkow ^35^ as an effective and applicable screening tool for cognitive disorders. The Thai version of MoCA was used to screen and monitor cognitive impairments in the clinical practice of neurocognitive disorders validated in the Thai setting by Tangwongchai, Phanasathit, Charernboon, Akkayagorn, Hemrungrojn, Phanthumchinda and Nasreddine ^33^. This test measures various cognitive domains, namely: visuospatial/executive, naming, attention, language, abstraction, delayed recall, and orientation. The total sum of all individual scores (out of 30 maximum possible points) represents the severity of cognitive impairment. The CERAD was developed in 1986 by the National Institute of Aging as a standardized, validated instrument to assess Alzheimer’s dementia. In the current study, we employ the Neuropsychological Assessment Battery (CERAD-NP), in a Thai validated translation ^7^. In this study, we used: the Word List Memory (WLM) to assess episodic memory and learning ability for new verbal information and immediate working memory; and Word List Recall, Delayed, True Recall (WLR) recognize to probe verbal episodic memory and the ability to recall. Verbal Fluency Test (VFT), assessing semantic memory or fluency, verbal productivity, language, and cognitive flexibility.

#### Psychological symptoms

Neuroticism traits were assessed with the Thai version of the 8-item subscale of the Five Factor Model standardized psychometric pool of items (IPIP-NEO; See http://ipip.ori.org) ^36^. The neuroticism subscale measures one’s tendency to experience negative emotions (“Have frequent mood swings”), and each item was rated on a 5-point Likert scale of 1 (Strongly Disagree) to 5 (Strongly Agree).

Stress symptoms were assessed using the 10-item *Perceived Stress Scale* (PSS) developed by Cohen and Williamson ^37^ and validated for use in a Thai setting by Wongpakaran and Wongpakaran ^38^. The scale assesses symptoms experienced during the last month preceding the survey. Each item is rated on a 5-point Likert-type scale from 0 (never) to 4 (very often). A higher score indicates greater distress. The Negative Life Events (NLE) is used to assess daily hassles ^39^. The NLE comprises 46 items covering the interpersonal stress in 10 domains, namely: household (6 items), problems with your work supervisor/employer (4 items), problems with parents (4 items), problems with spouse/partner (5 items), money hassles (4 items), problems with children (3 items), problems with friends (4 items), problems with other relatives (4 items), health hassles (4 items), problems with other workers (4 items), and work hassles (4 items). Participants indicate how much of a hassle each of the stressors was for them, using a 5-point Likert scale ranging from 0 (no hassle) to 4 (extreme hassle), and the scores were totalled to produce an overall hassles score. The NLE was back translated into Thai by Boonyamalik40.

Anxiety symptoms were assessed with the 20-item *State-Trait Anxiety Inventory* (STAI) developed by Spielberger, Gorsuch, Lushene, Vagg and Jacobs ^41^, and has been validated in the Thai setting by Iamsupasit and Phumivuthisarn ^42^. Each item is rated on a 4-point Likert scale ranging from 1 (not at all) to 4 (mostly). The total score ranges from 20-80 and higher scores are associated with greater feelings of anxiety. We also used the *anxiety subscale of the Hospital Anxiety and Depression Scale* (HADS-A) to assess severity of anxiety ^43^. The scale has been validated in the Thai setting by Nilchaikovit ^44^. This instrument consists of seven items rated on a four-point scale from 0 (not at all) to 3 (very often indeed), with five items reverse coded. The scores of the seven items were summed to create a scale that ranged from 0 to 21, with higher scores indicating more symptoms of anxiety. At the cutoff score ≥ 11, which was the best cutoff score, the sensitivity of the anxiety subscale of Thai HADS was 100% and the sensitivity was 86%.

The *Geriatric Depression Scale* (GDS) is a 30-item self-report measure designed to assess and screen depressive symptoms among older adults ^45^. It is a popular and widely used screening test to measure depression. The scale was validated and used with Thai elderly people for nearly three decades ^46^ (Train the Brain Forum Committee, 1994), with each response rated on a dichotomous scale of 0 and 1. A total score greater than 12 indicates depression, with higher scores suggesting a higher level of depression.

The pooled sensitivity and specificity of Thai GDS-30 are 82% and 76% ^47^. In addition, we used the *depression subscale of the Hospital Anxiety and Depression Scale* (HADS-D) ^43^. It has been validated in the Thai setting by Nilchaikovit ^44^. This instrument consists of seven items rated on a four-point scale from 0 (not at all) to 3 (very often indeed), with three items reverse coded. The scores of the seven items were summed to create a scale that ranged from 0 to 21, with higher scores indicating more symptoms of depression. At the cutoff score ≥ 11, which was the best cutoff score, the sensitivity of the depression subscale of Thai HADS was 85.71% respectively, while the specificity was 91.3%.

The original Adverse Childhood Experiences (ACE) study was used to identify childhood experiences of abuse or neglect at Kaiser Permanente from 1995 to 1997 with two waves of data collection ^48^. ACE were assessed using the Adverse Childhood Experiences Questionnaire in a Thai translation ^49^. This questionnaire consists of 28 items covering the traumatic experiences in childhood in 10 domains, namely: psychological abuse (2 items), physical abuse (2 items), sexual abuse (4 items), mental neglect (5 items), physical neglect (5 items), domestic violence (4 items), substance abuse in the family (2 items), family psychiatric illness (2 items), separation or divorce in the family (1 item), and family members in criminals (1 item) ^49^.

### Data analysis

Differences in continuous variables between groups were checked using analysis of variance (ANOVA). Analysis of contingency tables (the χ2-test) was used to determine the association between nominal variables. Correlations between two variables were assessed using Pearson’s product-moment correlation coefficients. Univariate general linear model (GLM) analysis was used to examine the relationships between diagnostic classifications and clinical and cognitive data after covarying for gender, age, and education. Subsequently, the estimated marginal means (SE) were computed from the GLM model after adjusting for the gender, age, and education variables. To examine pair-wise differences in group means, we employed the protected least significant difference. We performed multiple regression analysis to determine which test scores best predicted the clinical scores using a stepwise algorithm and to compute and display partial regression analysis of clinical data. For this analysis, we always confirmed multivariate normality, homoscedasticity, and the absence of collinearity and multicollinearity. The regression analyses’ results were always bootstrapped using 5,000 bootstrap samples, and the latter were reported if the findings were not concordant. Statistical tests were two-tailed and a p-value of 0.05 was used for statistical significance. IBM SPSS Windows version 29 was used for all statistical analyses.

Using the precision nomothetic approach ^46^, we aim to construct new phenotypes or phenotype dimensions by combining clinical data, rating scale scores and neurocognitive tests results. A two-step cluster analysis was performed considering categorical and continuous variables. The adequacy of the cluster solution was determined by evaluating the silhouette measure of cohesion and separation, which was deemed satisfactory if it exceeded a threshold of 0.3. Principal components analysis (PCA) was performed, and the Kaiser–Meier–Olkin (KMO) sample adequacy measure was used to assess factorability (sufficient when > 0.7). Moreover, when all loadings on the first factor were > 0.66, the variance explained by the first factor was > 50%, and Cronbach’s alpha performed on the variables was > 0.7, the first PC was regarded as a valid latent construct underpinning the variables.

Partial Least Squares path analysis (SmartPLS 4.0) was employed to check the paths between ACE, NLE, CERAD, MCI, and DSOA scores. Each variable was input as a single indicator (e.g., age and education; NLE health+money) or as reflective or formative constructs. We entered two reflective models: a) a first factor extracted from neuroticism, HADS-D, HADS-A, PSS, STAI, and GDS (thus reflecting the DSOA construct), and b) a second from MMSE, MOCA and Petersen item 1 (thus reflecting MCI). Moreover, we entered two formative models: a) a first based on emotional abuse, emotional neglect, and physical neglect (a composite of ACE); and b) a second formative model based on three CERAD test results, namely VFT, WLM and WLR. We conducted complete PLS analysis using 5,000 bootstrap samples only when the outer and inner models complied with quality data, namely the factors have an adequate average variance extracted (AVE) > 0.5, and rho a > 0.8; all LV loadings are > 0.66 at p < 0.001; the model fit as assessed with SRMR is adequate (SRMR < 0.08); and Confirmatory Tetrad analysis (CTA) shows that the LV is not mis specified as a reflective model.

## Results

### Results of factor analyses

We employed principal component analysis (PCA) to examine whether latent vectors could be extracted from neuroticism (IPIP-NEO subdomain), PSS, HADS-A, HADS-D, STAI, TGDS. Table 1 shows the results of these PC analyses. The first PCA showed a sufficient factorability of the correlation matrix and the first factor explained 60.14% of the variance and all factor loadings were > 0.670 with an adequate Cronbach’s alpha value. This first PC, therefore, was dubbed the “Distress Symptoms of Old Age” or “DSOA”. We could also extract a single latent vector from the MoCA, TMSE, CERAD-WLR, and CERAD WLM with adequate KMO and explained variance values, and a sufficient Cronbach’s alpha value (see Table 1). Because this PC reflect a mild cognitive dysfunction, it was labelled as mCoDy. The VFT and CDR scores did not load highly on this PC (<0.5) and were, therefore, deleted from the PCA #2 model. Nevertheless, we were able to extract one validated PC from the MoCa, MMSE and CDR scores (see Table 1). Because this PC comprises the CDR score (0: control, 0.5: aMCI) we called this PC “quantitative MCI (qMCI).” As such, we computed two different cognitive impairment scores, a first focusing on mild memory impairments and a second on a mild, more generalized cognitive decline. We were unable to combine any of the cognitive test results with the affective, stress, anxiety, or neuroticism scores. **Figure 1** shows that there is no significant association between the DSOA and mCoDy PC scores.

**Table 1.**
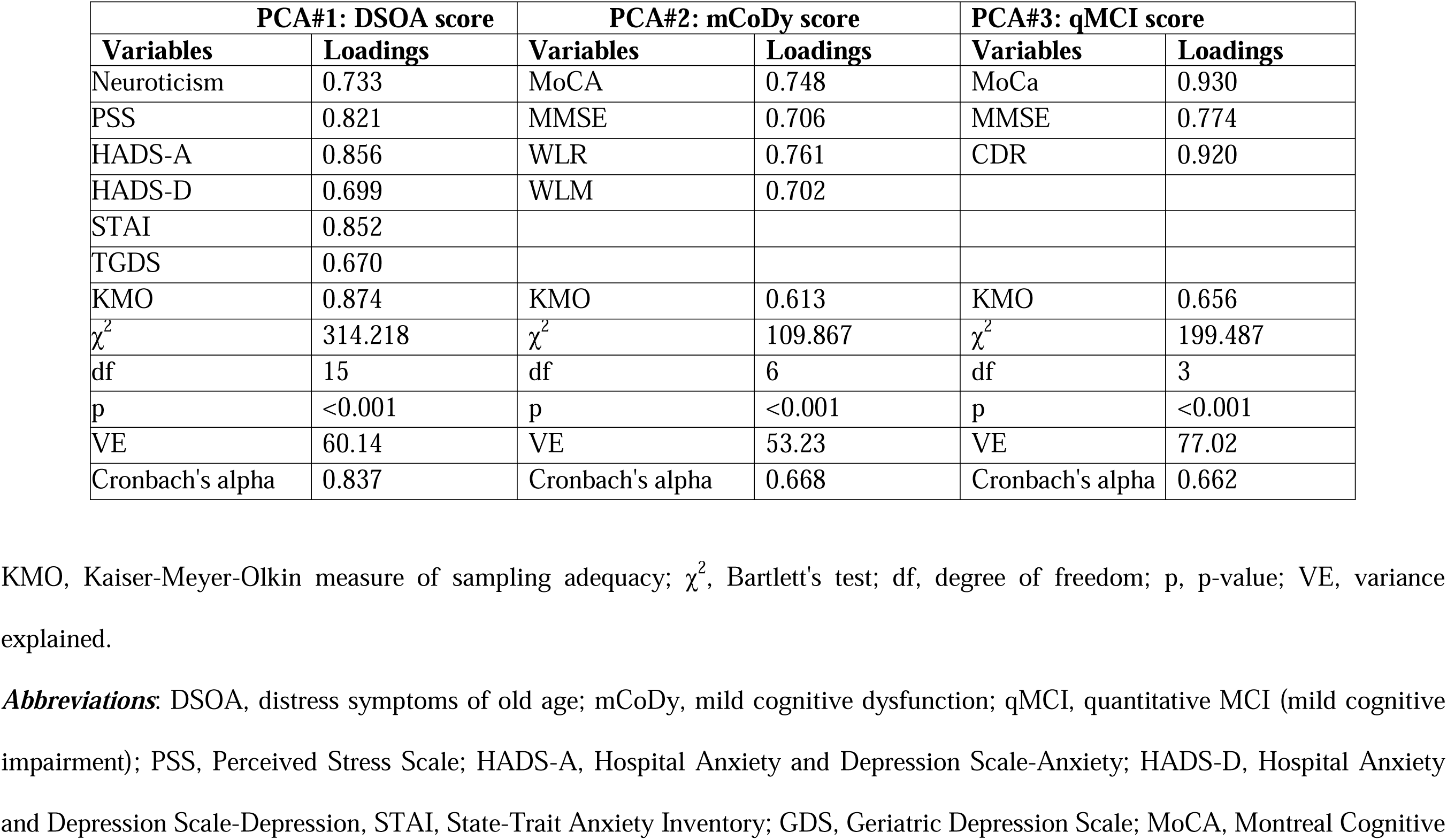
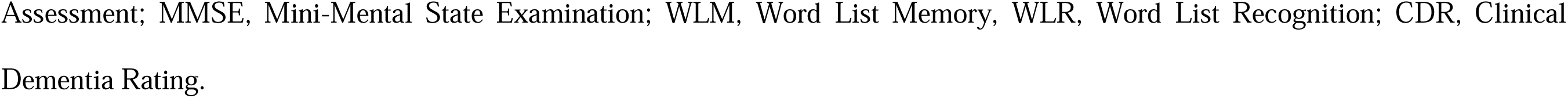
Results of principal component analyses (PCA)

| PCA#1: DSOA score |  | PCA#2: mCoDy score |  | PCA#3: qMCI score |  |
| --- | --- | --- | --- | --- | --- |
| Variables | Loadings | Variables | Loadings | Variables | Loadings |
| Neuroticism | 0.733 | MoCA | 0.748 | MoCa | 0.930 |
| PSS | 0.821 | MMSE | 0.706 | MMSE | 0.774 |
| HADS-A | 0.856 | WLR | 0.761 | CDR | 0.920 |
| HADS-D | 0.699 | WLM | 0.702 |  |  |
| STAI | 0.852 |  |  |  |  |
| TGDS | 0.670 |  |  |  |  |
| KMO | 0.874 | KMO | 0.613 | KMO | 0.656 |
| $\chi^2$ | 314.218 | $\chi^2$ | 109.867 | $\chi^2$ | 199.487 |
| df | 15 | df | 6 | df | 3 |
| p | <0.001 | p | <0.001 | p | <0.001 |
| VE | 60.14 | VE | 53.23 | VE | 77.02 |
| Cronbach's alpha | 0.837 | Cronbach's alpha | 0.668 | Cronbach's alpha | 0.662 |

KMO, Kaiser-Meyer-Olkin measure of sampling adequacy; $\chi^2$ , Bartlett's test; df, degree of freedom; p, p-value; VE, variance explained.
**Abbreviations:** DSOA, distress symptoms of old age; mCoDy, mild cognitive dysfunction; qMCI, quantitative MCI (mild cognitive impairment); PSS, Perceived Stress Scale; HADS-A, Hospital Anxiety and Depression Scale-Anxiety; HADS-D, Hospital Anxiety and Depression Scale-Depression, STAI, State-Trait Anxiety Inventory; GDS, Geriatric Depression Scale; MoCA, Montreal Cognitive
Assessment; MMSE, Mini-Mental State Examination; WLM, Word List Memory, WLR, Word List Recognition; CDR, Clinical Dementia Rating.

**Figure 1.**
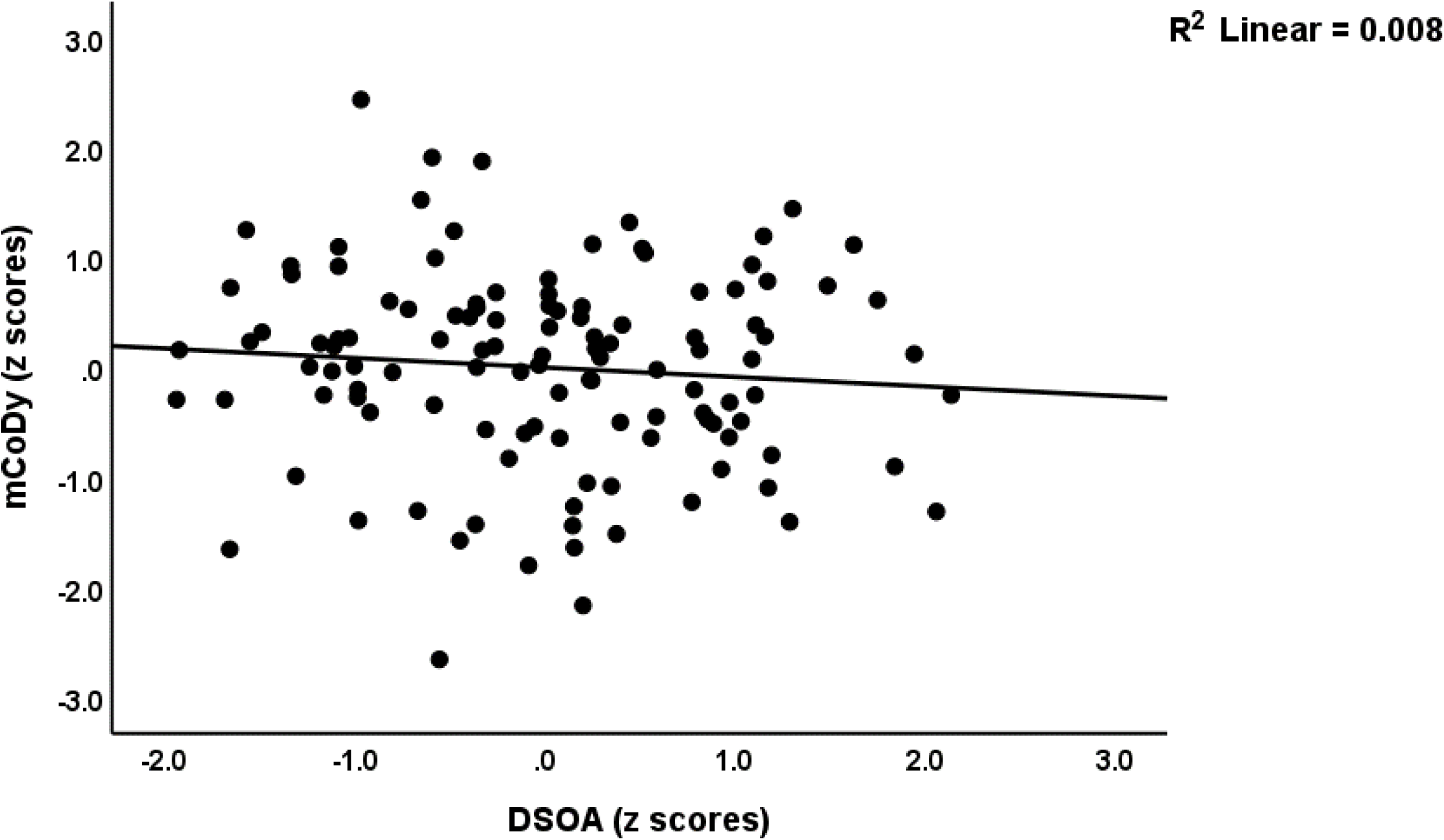
Partial regression plot of the mild cognitive dysfunction (mCoDy) score on the distress symptoms of old age (DSOA) score (p=0.336, after adjustment for sex, age and education).

### Socio-demographic and Clinical Data

A two-step clustering analysis with automatic determination of the number of clusters and Schwarz’s Bayesian Criterion (BIC) was conducted to determine if genuine clusters of subjects with aMCI or affective symptoms could be identified. Toward this end we performed two-step cluster analysis using the following variables: clinical diagnosis of aMCI, neuroticism, PSS, STAI, HADS-A, HAD-D, TGDS, WLM, WLR, VFT, MoCa and MMSE scores. Automatic determination of the number of clusters showed that three clusters could be retrieved from the data set with a silhouette measure of cohesion and separation of 0.4, indicating a fair cluster solution. There were 37 subjects in cluster 1, 31 in cluster 2, and 52 in cluster 3.

**Table 2** shows the features of the three groups. Cluster 3 individuals were significantly older and has significantly lower education years than cluster 1 and cluster 2 individuals. Of the 59 healthy controls included in the study, the cluster analyses allocated 34 to the control group, 22 to cluster 2 and 3 to cluster 3. Of the 61 subjects with aMCI included in the study, the cluster analysis allocated 49 to cluster 3, 9 to cluster 2 and 3 to cluster 1 (see Table 2). From the demographics and clinical features of the three study groups, no significant differences were found in sex, or marital status distribution between the three groups. The DSOA score are significantly different between the three cluster and increased from cluster 1 to cluster 3 to cluster 2. The mCoDy scores were significantly greater in cluster 3 than in cluster 1 and 2; there were no significant differences between cluster 1 and 2 in the mCoDy scores. Therefore, we have labelled cluster 2 as a DSOA cluster, cluster 2 as a mCoDy cluster, and cluster 1 as the control cluster. The differences between these three clusters in DSOA (F-49.30, df=2/113, p<0.001) and mCoDy (F=78.01, df=2/113, p<0.001) score as well as the intergroup differences remained significant after covarying for age, sex, and education. Neuroticism, PSS, HADS-A, HADS-D, and STAI scores were significantly different between the three groups and increased from controls to mCoDy to DSOA. VFT, WLM, WLR and MMSE scores were significantly lower in mCoDy participants than in controls and DSOA subjects. The MoCA score was significantly different between the three clusters as decreased from controls to the DSOA to mCoDy clusters.

**Table 2.**
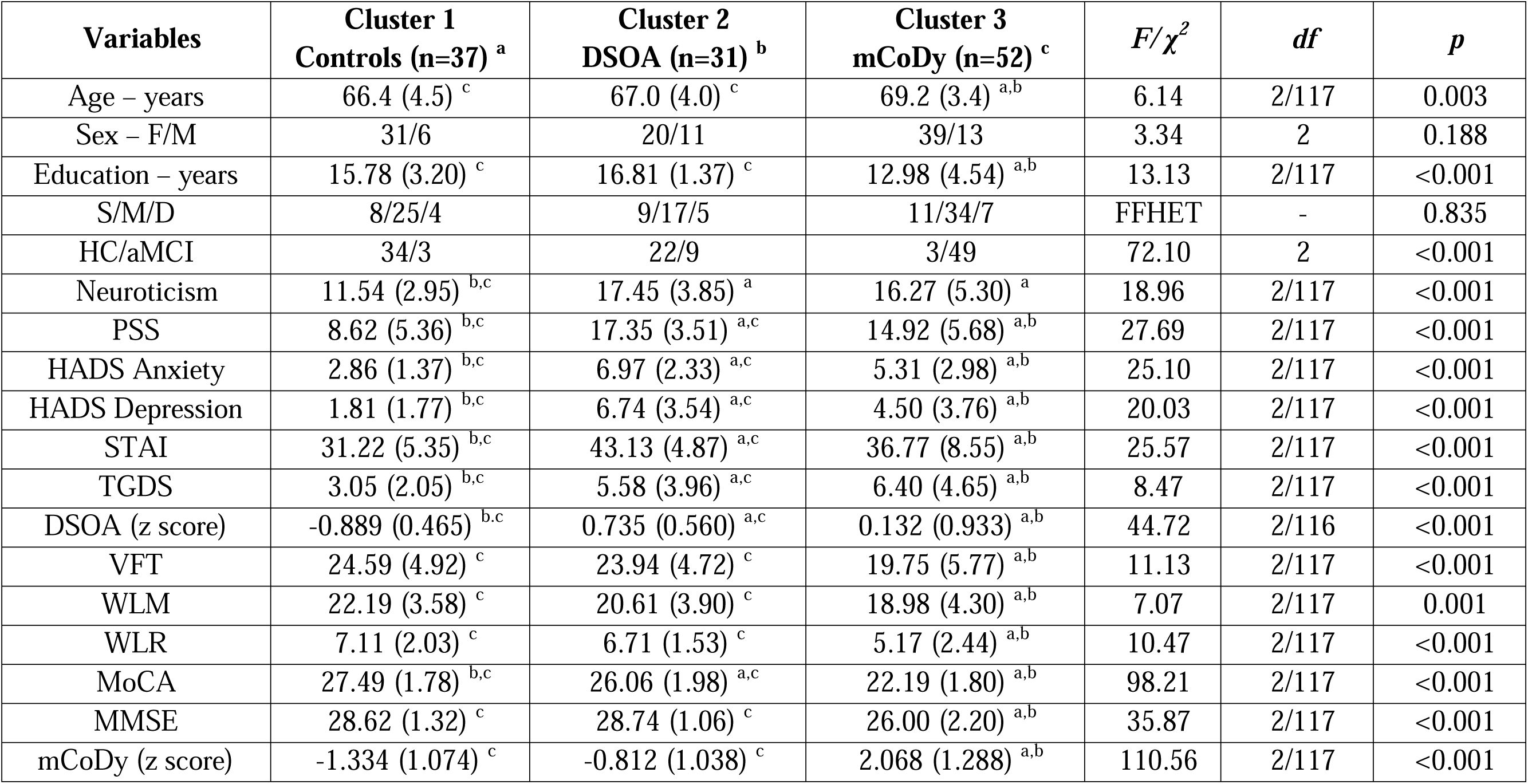
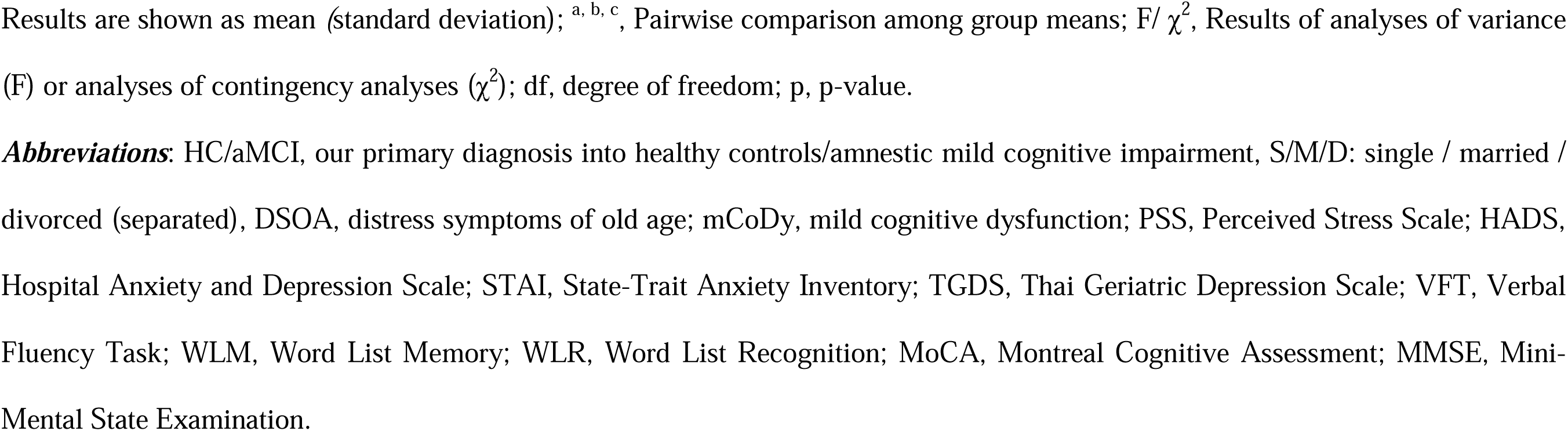
Socio-demographic and clinical data in three clusters of old adults: those with distress symptoms of old age (DSOA), those with mild cognitive dysfunction (mCoDy), and those without mCoDy/DSOA (controls).

| **Variables** | **Cluster 1**<br>**Controls (n=37) <sup>a</sup>** | **Cluster 2**<br>**DSOA (n=31) <sup>b</sup>** | **Cluster 3**<br>**mCoDy (n=52) <sup>c</sup>** | ***F/χ<sup>2</sup>*** | ***df*** | ***p*** |
| --- | --- | --- | --- | --- | --- | --- |
| Age – years | 66.4 (4.5) <sup>c</sup> | 67.0 (4.0) <sup>c</sup> | 69.2 (3.4) <sup>a,b</sup> | 6.14 | 2/117 | 0.003 |
| Sex – F/M | 31/6 | 20/11 | 39/13 | 3.34 | 2 | 0.188 |
| Education – years | 15.78 (3.20) <sup>c</sup> | 16.81 (1.37) <sup>c</sup> | 12.98 (4.54) <sup>a,b</sup> | 13.13 | 2/117 | <0.001 |
| S/M/D | 8/25/4 | 9/17/5 | 11/34/7 | FFHET | - | 0.835 |
| HC/aMCI | 34/3 | 22/9 | 3/49 | 72.10 | 2 | <0.001 |
| Neuroticism | 11.54 (2.95) <sup>b,c</sup> | 17.45 (3.85) <sup>a</sup> | 16.27 (5.30) <sup>a</sup> | 18.96 | 2/117 | <0.001 |
| PSS | 8.62 (5.36) <sup>b,c</sup> | 17.35 (3.51) <sup>a,c</sup> | 14.92 (5.68) <sup>a,b</sup> | 27.69 | 2/117 | <0.001 |
| HADS Anxiety | 2.86 (1.37) <sup>b,c</sup> | 6.97 (2.33) <sup>a,c</sup> | 5.31 (2.98) <sup>a,b</sup> | 25.10 | 2/117 | <0.001 |
| HADS Depression | 1.81 (1.77) <sup>b,c</sup> | 6.74 (3.54) <sup>a,c</sup> | 4.50 (3.76) <sup>a,b</sup> | 20.03 | 2/117 | <0.001 |
| STAI | 31.22 (5.35) <sup>b,c</sup> | 43.13 (4.87) <sup>a,c</sup> | 36.77 (8.55) <sup>a,b</sup> | 25.57 | 2/117 | <0.001 |
| TGDS | 3.05 (2.05) <sup>b,c</sup> | 5.58 (3.96) <sup>a,c</sup> | 6.40 (4.65) <sup>a,b</sup> | 8.47 | 2/117 | <0.001 |
| DSOA (z score) | -0.889 (0.465) <sup>b,c</sup> | 0.735 (0.560) <sup>a,c</sup> | 0.132 (0.933) <sup>a,b</sup> | 44.72 | 2/116 | <0.001 |
| VFT | 24.59 (4.92) <sup>c</sup> | 23.94 (4.72) <sup>c</sup> | 19.75 (5.77) <sup>a,b</sup> | 11.13 | 2/117 | <0.001 |
| WLM | 22.19 (3.58) <sup>c</sup> | 20.61 (3.90) <sup>c</sup> | 18.98 (4.30) <sup>a,b</sup> | 7.07 | 2/117 | 0.001 |
| WLR | 7.11 (2.03) <sup>c</sup> | 6.71 (1.53) <sup>c</sup> | 5.17 (2.44) <sup>a,b</sup> | 10.47 | 2/117 | <0.001 |
| MoCA | 27.49 (1.78) <sup>b,c</sup> | 26.06 (1.98) <sup>a,c</sup> | 22.19 (1.80) <sup>a,b</sup> | 98.21 | 2/117 | <0.001 |
| MMSE | 28.62 (1.32) <sup>c</sup> | 28.74 (1.06) <sup>c</sup> | 26.00 (2.20) <sup>a,b</sup> | 35.87 | 2/117 | <0.001 |
| mCoDy (z score) | -1.334 (1.074) <sup>c</sup> | -0.812 (1.038) <sup>c</sup> | 2.068 (1.288) <sup>a,b</sup> | 110.56 | 2/117 | <0.001 |

Results are shown as mean (standard deviation); <sup>a, b, c</sup>, Pairwise comparison among group means; F/ $\chi^2$ , Results of analyses of variance (F) or analyses of contingency analyses ( $\chi^2$ ); df, degree of freedom; p, p-value.
**Abbreviations:** HC/aMCI, our primary diagnosis into healthy controls/amnestic mild cognitive impairment, S/M/D: single / married / divorced (separated), DSOA, distress symptoms of old age; mCoDy, mild cognitive dysfunction; PSS, Perceived Stress Scale; HADS, Hospital Anxiety and Depression Scale; STAI, State-Trait Anxiety Inventory; TGDS, Thai Geriatric Depression Scale; VFT, Verbal Fluency Task; WLM, Word List Memory; WLR, Word List Recognition; MoCA, Montreal Cognitive Assessment; MMSE, Mini- Mental State Examination.

### Differences in ACE and NLE scores between the diagnostic groups

**Table 3** shows that the ACE and NLE scores in the three clusters. While there were no significant differences in the 5 ACE that were examined in this study, subjects with DSOA showed significant increased ACE scores as compared with controls. Although there was no overall significance between the three clusters in total NLE score, there was a trend towards higher NLE scores in the DSOA cluster as compared with the control cluster, while subjects in the mCoDy cluster took up an intermediate position. Examining the separate NLE items showed that the sum of money + health NLE scores was significantly higher in both the DSOA and mCoDy subjects as compared with controls.

**Table 3.** Adverse childhood experiences (ACE) and negative life events (NLE) in three clusters of old adults: those with distress symptoms of old age (DSOA), those with mild cognitive dysfunction (mCoDy), and those without mCoDy/DSOA (controls).

| Variables | Controls (n=37)<sup>a</sup> | DSOA (n=31)<sup>b</sup> | mCoDy (n=52)<sup>c</sup> | *F* | *df* | *p* |
| --- | --- | --- | --- | --- | --- | --- |
| ACE |  |  |  |  |  |  |
| Emotional abuse | 0.27 (0.56) | 0.42 (0.67) | 0.35 (0.65) | 0.47 | 2/117 | 0.624 |
| Physical abuse | 0.24 (0.49) | 0.35 (0.55) | 0.17 (0.38) | 1.48 | 2/117 | 0.232 |
| Sexual abuse | 0.03 (0.16) | 0.32 (0.94) | 0.10 (0.49) | 2.33 | 2/117 | 0.102 |
| Emotional neglect | 9.65 (3.67) | 11.55 (4.49) | 11.13 (4.35) | 2.05 | 2/117 | 0.133 |
| Physical neglect | 7.19 (2.08)<sup>c</sup> | 8.10 (4.50) | 8.60 (2.97)<sup>a</sup> | 3.18 | 2/117 | 0.045 |
| Sum 5 ACEs (z scores) | -0.310 (0.991)<sup>b</sup> | 0.318 (1.001)<sup>a</sup> | 0.035 (0.968) | 3.49 | 2/117 | 0.034 |
| Sum neglect | 16.84 5.06 | 19.65 6.14 | 19.73 6.29 | 3.01 | 2/117 | 0.053 |
| NLE |  |  |  |  |  |  |
| Total NLE * | 12.89 (14.21)<sup>b</sup> | 19.30 (13.73)<sup>a</sup> | 14.92 (13.34) | 2.72 | 2/115 | 0.070 |
| NLE money+health | 1.70 (2.72)<sup>b,c</sup> | 3.70 (3.48)<sup>a</sup> | 3.13 (3.27)<sup>a</sup> | 5.95 | 2/116 | 0.003 |

Results are shown as mean (SD); <sup>a, b, c</sup>, Pairwise comparison among group means; *F*, Results of analyses of variance; *df*, degree of freedom; *p*, p-value. **Abbreviations:** HC, Healthy Control; DSOA, distress symptoms of old age; mCoDy, mild cognitive dysfunction; ACE, Adverse childhood experiences; NLE, Negative Life Events.

### Prediction of the DSOA and mCoDy scores

We performed different multiple regression analyses using the DSOA and mCoDy scores as dependent variables and age, sex, education, NLE, and ACE data as explanatory variables (with and without Petersen item 1) See **Table 4**. Regression #1 shows that 32.8% of the variance in DSOA score could be explained by NLE health+money, ACE neglect, and education (all three positively associated). **Figure 2** shows the partial regression plot (after considering the effects of age, sex, education) of the DSOA score on NLE health+money showing a significant positive association between both variables. **Figure 3** shows the partial regression plot of the DSOA score on ACE neglect (after adjusting for age, sex, and education). **Figure 4** shows the partial regression plot of the mCoDy score on education years. Table 4, regression #2 shows the prediction of the DSOA score was somewhat improved after considering the effects of Petersen, item 1. Thus, 37.9% of the variance in DSOA score could be explained by NLE health+money, ACE neglect, education, and Petersen_1 (all positively associated). The latter is important as it shows that subjective feelings of cognitive impairment, but not objective measurements of cognitive impairment, may increase DSOA.

**Table 4.**
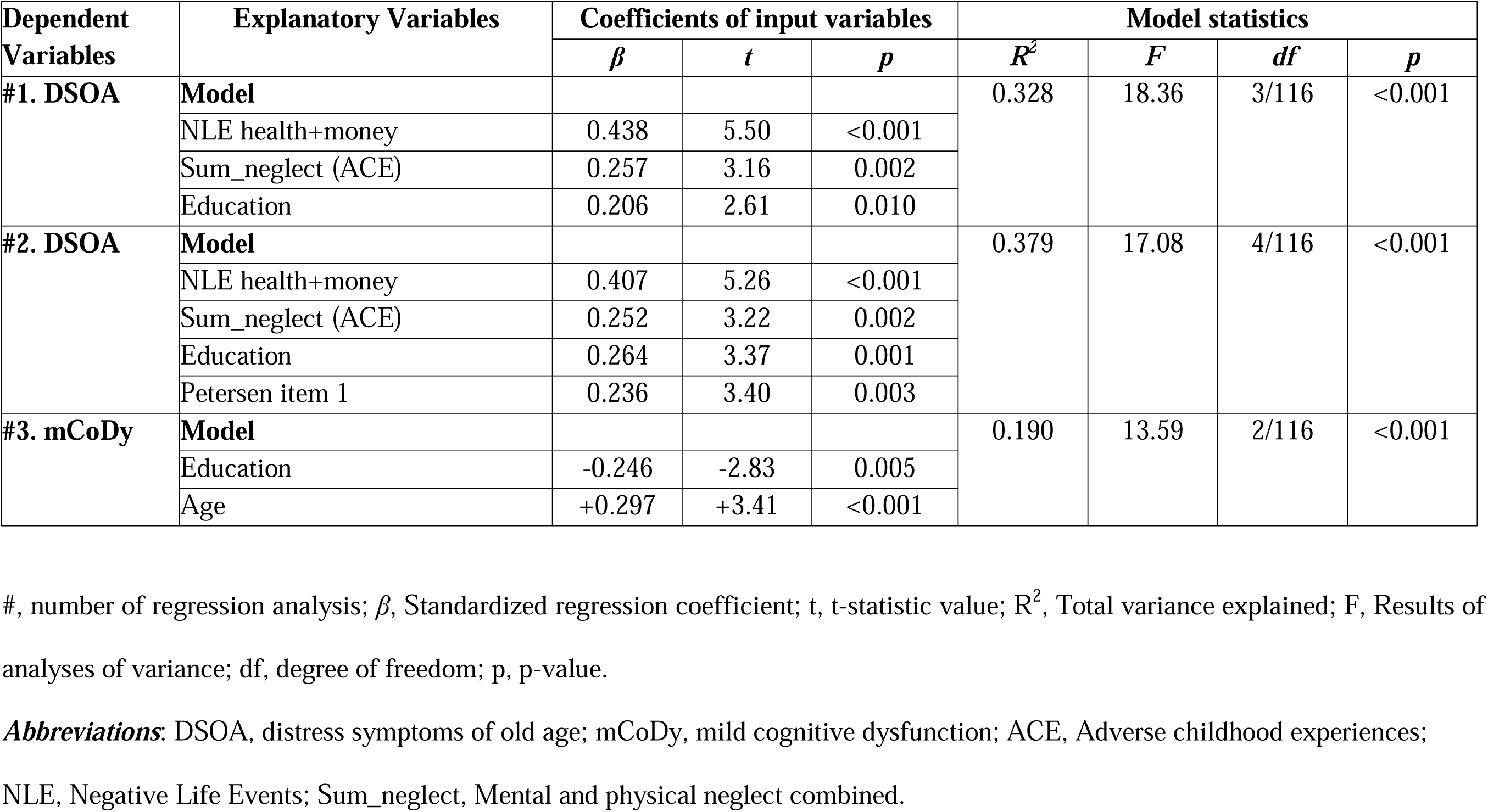
Results of multiple regression analyses with distress symptoms of old age (DSOA) and mild cognitive dysfunction (mCoDy) scores as dependent variables

| Dependent Variables | Explanatory Variables | Coefficients of input variables |  |  | Model statistics |  |  |  |
| --- | --- | --- | --- | --- | --- | --- | --- | --- |
| | | $\beta$ | $t$ | $p$ | $R^2$ | $F$ | $df$ | $p$ |
| #1. DSOA | **Model** |  |  |  | 0.328 | 18.36 | 3/116 | <0.001 |
|  | NLE health+money | 0.438 | 5.50 | <0.001 |  |  |  |  |
|  | Sum_neglect (ACE) | 0.257 | 3.16 | 0.002 |  |  |  |  |
|  | Education | 0.206 | 2.61 | 0.010 |  |  |  |  |
| #2. DSOA | **Model** |  |  |  | 0.379 | 17.08 | 4/116 | <0.001 |
|  | NLE health+money | 0.407 | 5.26 | <0.001 |  |  |  |  |
|  | Sum_neglect (ACE) | 0.252 | 3.22 | 0.002 |  |  |  |  |
|  | Education | 0.264 | 3.37 | 0.001 |  |  |  |  |
|  | Petersen item 1 | 0.236 | 3.40 | 0.003 |  |  |  |  |
| #3. mCoDy | **Model** |  |  |  | 0.190 | 13.59 | 2/116 | <0.001 |
|  | Education | -0.246 | -2.83 | 0.005 |  |  |  |  |
|  | Age | +0.297 | +3.41 | <0.001 |  |  |  |  |

#, number of regression analysis; $\beta$ , Standardized regression coefficient; $t$ , t-statistic value; $R^2$ , Total variance explained; $F$ , Results of analyses of variance; $df$ , degree of freedom; $p$ , p-value.
**Abbreviations:** DSOA, distress symptoms of old age; mCoDy, mild cognitive dysfunction; ACE, Adverse childhood experiences; NLE, Negative Life Events; Sum\_neglect, Mental and physical neglect combined.

**Figure 2.**
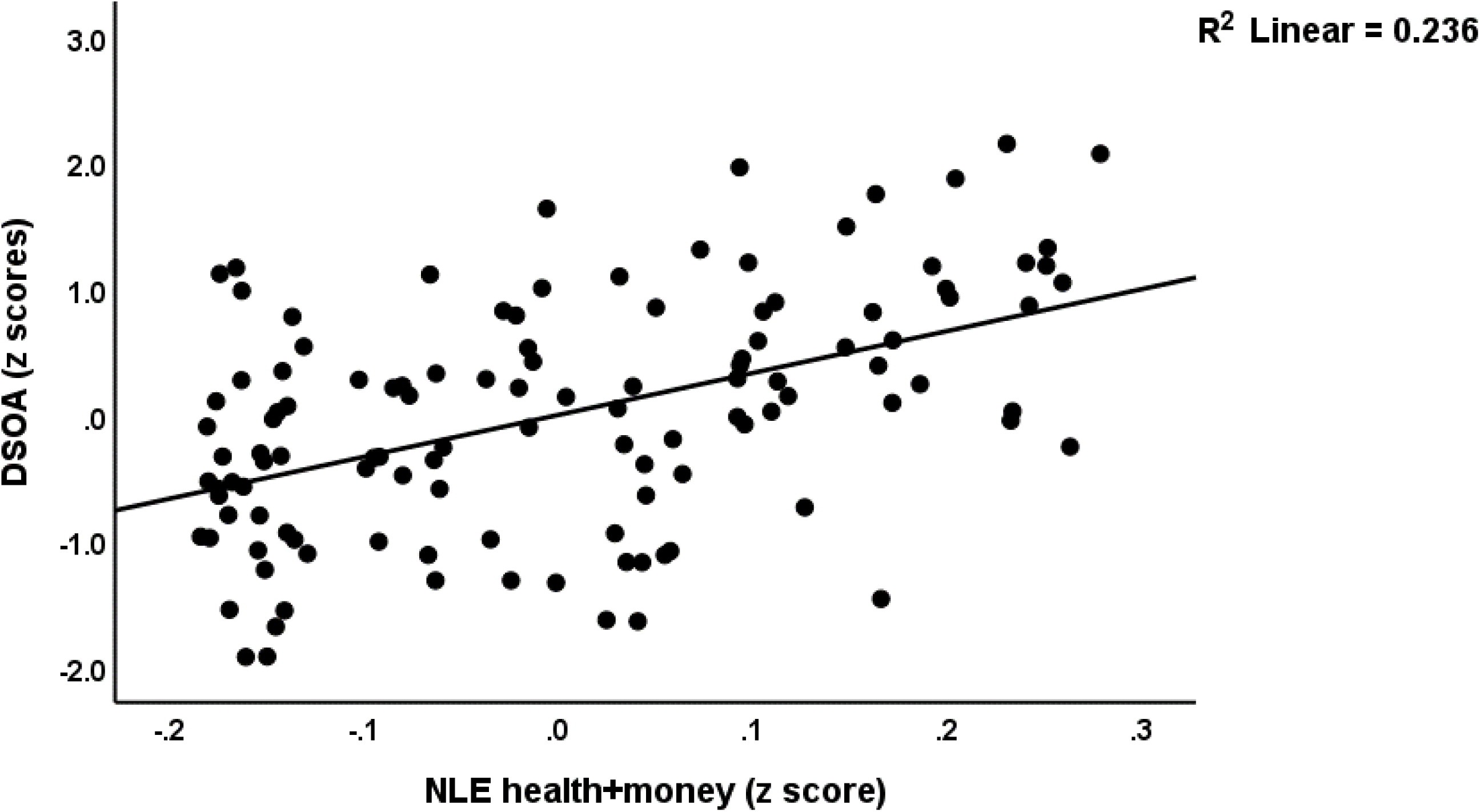
Partial regression plot of the distress symptoms of old age (DSOA) score on negative life events (health + money) (p<0.001).

**Figure 3.**
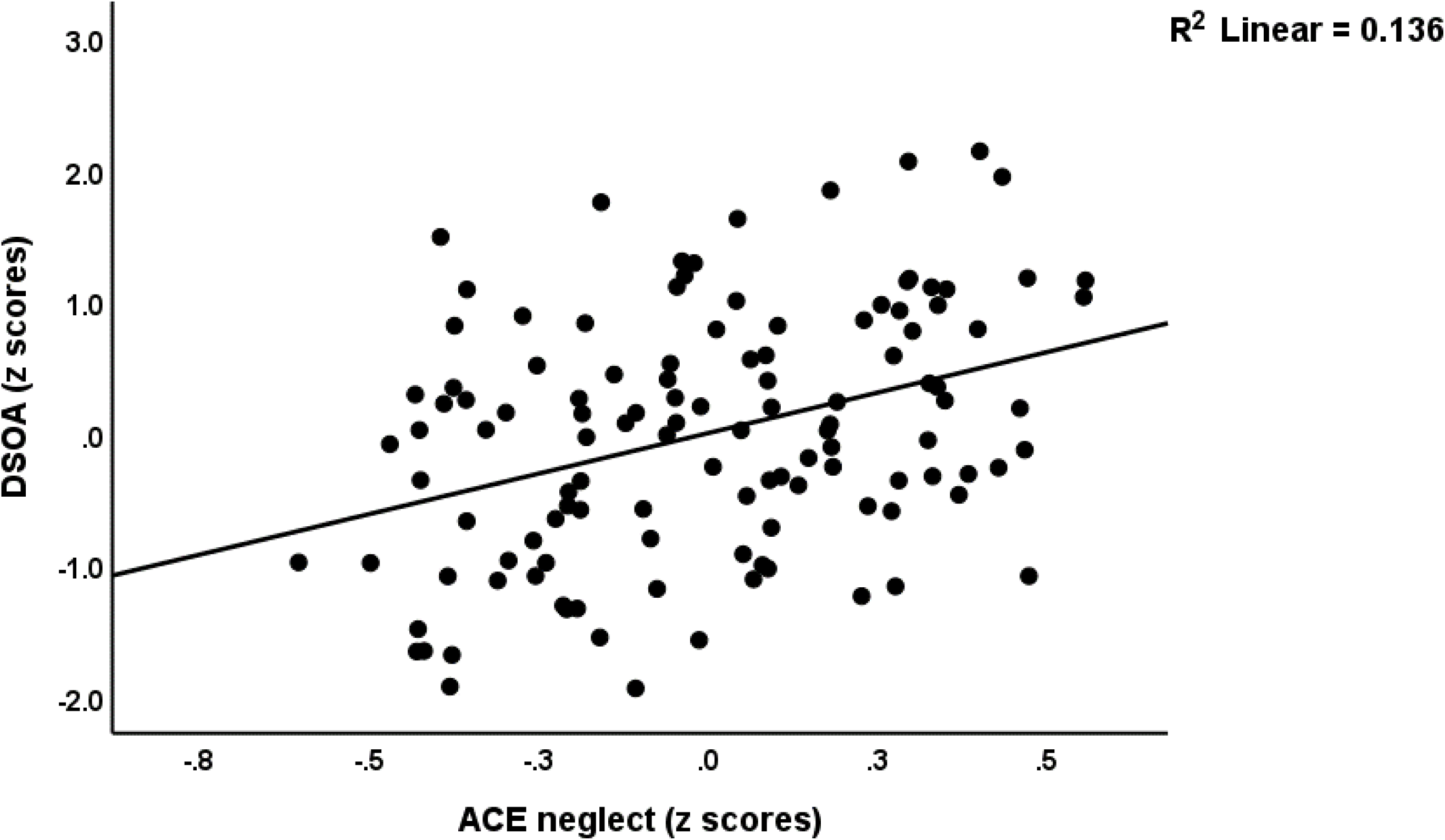
Partial regression plot of the distress symptoms of old age (DSOA) score on ACE physical + emotional neglect (p=0.002).

**Figure 4.**
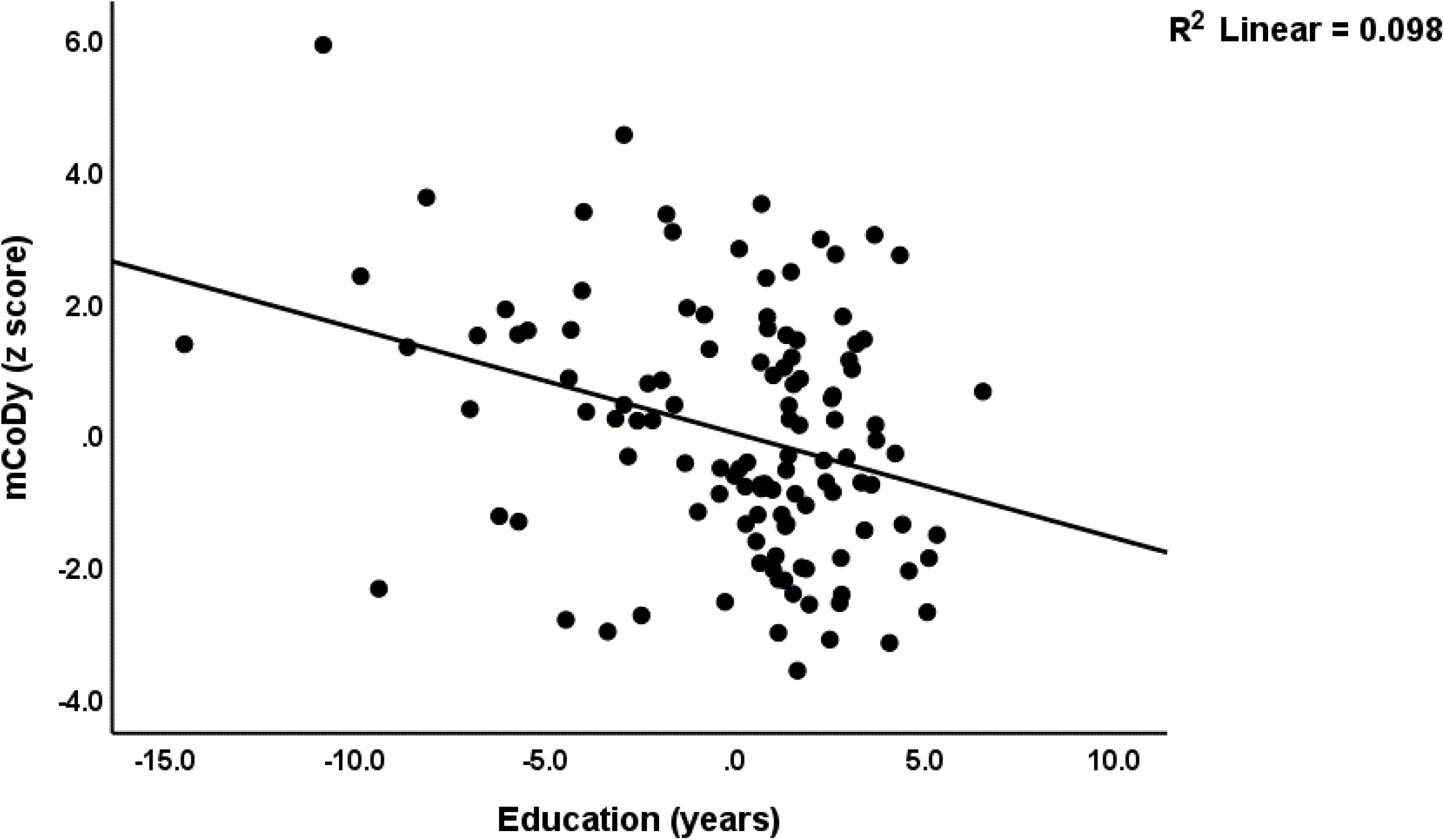
Partial regression plot of the mild cognitive dysfuction score (mCoDy) on education years (p<0.001).

Table 4, regression #3 shows that 19.0% of the variance in the mCoDy score was explained by the combined effects of age and education. There were no significant effects of the NLE or ACE variables on the mCoDy score.

### Results of PLS-path analysis

**Figure 5** shows the final PLS model after feature reduction (e.g., sex was excluded as there were no significant effects). We entered Petersen item 1, qMCI (a proxy of the MCI diagnosis), and the CERAD tests results as predictors of DSOA to examine if the latter was predicted by any of these clinical cognitive data. We entered two latent vectors, one reflecting clinical MCI symptom severity (extracted from CRC, MoCA and MMSE scores), and a second reflecting DSOA (extracted from HADS_A, HADS_D, neuroticism, PSS, STAI, and TGDS scores). ACE and CERAD were entered as two formative models using emotional abuse and neglect, and physical neglect, and VFT, WLR, and WLM, respectively. All other variables were entered as simple indicators. The model fit was more than adequate with a SRMR of 0.055. We observed adequate convergence and construct reliability validity values for (a) the DSOA latent factor with AVE = 0.598, and rho_a = 0.871, while all loadings were > 0.680 at p < 0.001; and (b) the MCI factor with AVE = 0.770, and rho_a = 0.864, while the loadings of the MCI construct were > 0.771. Both latent vectors were not mis-specified as reflective models (results of CTA). Complete PLS analysis, performed using 5,000 bootstraps, showed that 40.9% of the variance in the DSOA score was explained by the clinical Petersen item 1, ACE, NLE health + money, and education, whilst 28.9% of the variance in the clinical MCI score was explained by CERAD and education years. ACE and NLE did not have any effects on CERAD or the clinical MCI scores. Total effect analyses showed that ACE (t=3.65, p<0.001), NLE health+money (t=5.50, p<0.001) were the most important predictors of the DSOA score, followed at a distance by the clinical MCI score (t=2.38, p=0.018) and education (t=2.65, p=0.008).

**Figure 5.**
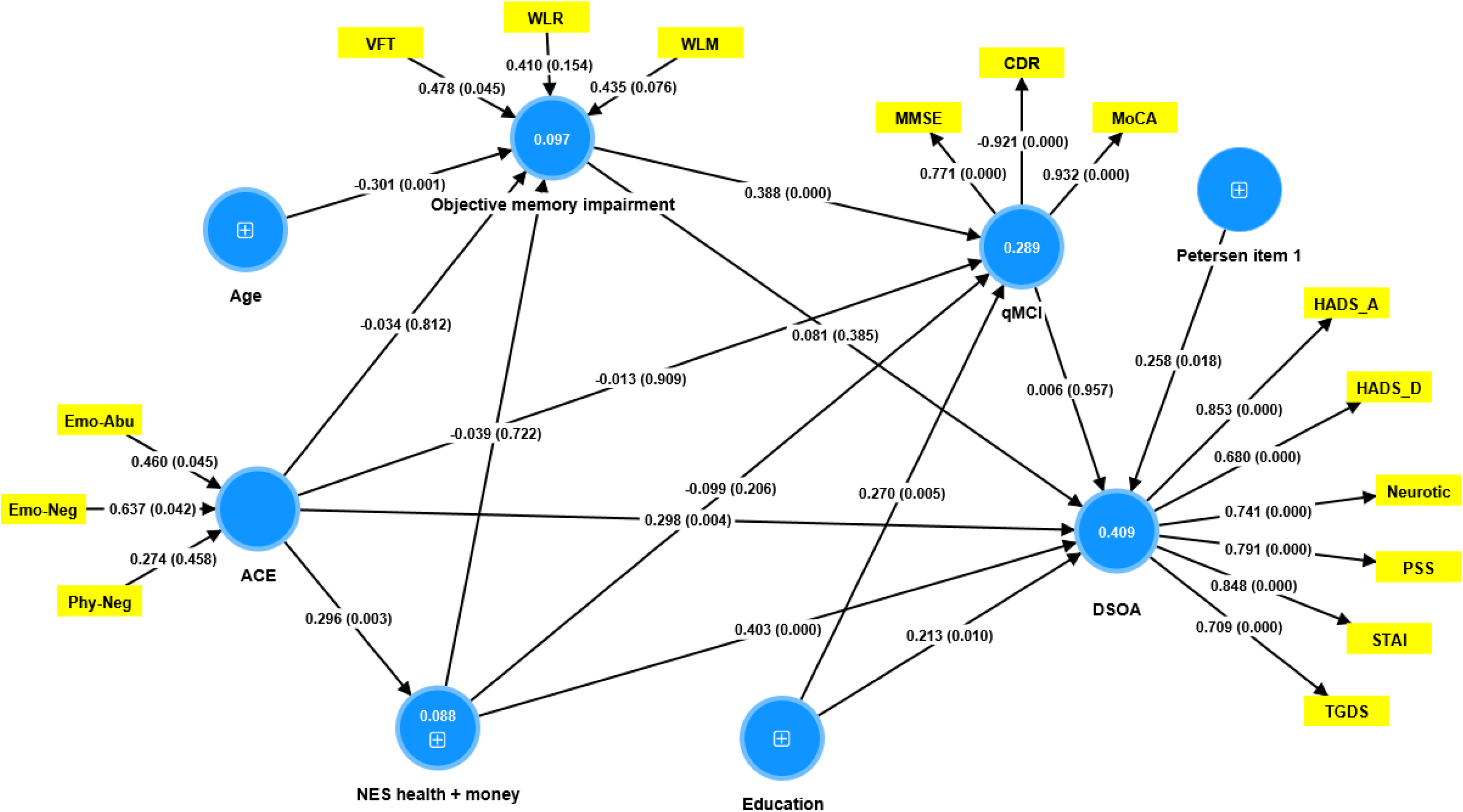
Results of partial least squares analysis. Entered in the analysis are two reflective constructs, namely a) DSOA (distress symptoms of old age) conceptualized as a factor extracted from PSS (Perceived Stress Scale), HADS (Hospital Anxiety and Depression Scale), STAI (State-Trait Anxiety Inventory) and TGDS (Thai Geriatric Depression Scale) scores; and b) qMCI (a quantitative mild cognitive impairment score) conceptualized as a factor extracted from CDR (modified Clinical Dementia Rating), MoCA (Montreal Cognitive Assessment) and MMSE (Mini-Mental State Examination) scores. This model also entered two formative constructs, namely a) objective memory impairment comprising VFT (verbal fluency test), WLM (World List Memory) and WLR (Word List Recognition) scores; and b) ACE (adverse childhood experiences) comprising Em-Abu (emotional abuse), Emo-neglect (emotional neglect), and Phys-neglect (physical neglect). The other variables are entered as single indicators including NES health+money (negative life events due to health and money problems). Shown are the pathway coefficients (with exact p-value). Figures within the circles denote explained variance. Reflective models are specified using loadings (with p-values) on the factors, and the formative models by the weights (p values) of the indicators.

## Discussion

### Presence of a DSOA dimension in older adults

The primary finding of this research is that we were able to extract one validated principal component or factor from distress, anxiety, depression, and neuroticism scores in older adults without major depression and ADL dysfunctions. The positive contributors to this principal component encompass symptoms of distress (PSS), anxiety (HADS-A and STAI), depression (HADS-D and TGDS) and neuroticism. Earlier research conducted by Jirakran, Vasupanrajit, Tunvirachaisakul and Maes ^23^ revealed that one principal component or factor could be extracted from anxiety, depression and neuroticism scores in adults with major depression. Neuroticism has been linked with mood and anxiety symptoms in both clinical ^50^ and non-clinical ^51–53^ research cohorts, as well as in cross-sectional investigations ^54,55^. Individual with increased neuroticism suffer from mood swings, feelings of guilt, anxiety, and depression, and they cope poorly with psychological stressors ^23^.

Distress in old age is a common phenomenon and is often linked with a range of physical and psychological symptoms such as stress, anxiety, melancholy, and neuroticism ^56^. However, these symptoms do not always indicate major depression, which causes significant distress and impedes daily functioning ^57^. Although depression is a frequent psychiatric disorder in a geriatric population and a substantial risk factor for disability and mortality, it is inaccurately diagnosed in nearly half of all instances ^57^. Another study showed that the levels of stress, anxiety, depression, and neuroticism are correlated with symptoms of geriatric discomfort, such as melancholy, fatigue, ambiguous physical complaints, and withdrawal from activities or social circles ^58^.

Consequently, the principal component derived here from distress, affective, and neuroticism scores in older adults without clinical depression and ADL dysfunctions reflects more a distress or psychological strain score rather than a “clinical depression” score. Therefore, we labelled this score the “Distress Symptoms of Old Age” (DSOA) score. As such, this is a new construct indicating that mild subjective distress symptoms may occur in part of the older adults independently from major depression. The subclinical symptom domain differs significantly from the behavioral and psychological symptoms of dementia (BPSD) which may occur in patients with dementia and some subjects with MCI ^56^. The latter symptoms domain encompass delusions, hallucinations, agitation/aggression, depression/dysphoria, anxiety, euphoria, apathy/indifference, disinhibition, irritability/lability, aberrant motor behaviour, nocturnal behavioral disturbances, and irregularities in appetite and eating ^56^. It should be added that this study excluded subjects with psychosis, either hallucinations or delusions, agitation, euphoria, or aberrant behaviours and thus excluded not only patients with major depression but also subjects with BPSD. This is important as some patients with aMCI may show BPSD ^56^.

All in all, DSOA is a subclinical symptom domain that emphasizes the presence of distress, mild affective symptoms, and increased neuroticism scores and thus reflects mild psychological strain rather than major depression or BPSD.

### Presence of mild cognitive dysfunction (mCoDy) in older adults

The second major finding of this study is that we were able to extract one cognitive dimension (principal component or factor) comprising the MoCa, MMSE, WLM, and WLR scores and indicating mild cognitive dysfunction (mCoDy). Increasing mCoDy scores suggest a decline in specific cognitive functions, particularly episodic memory, as well as a more generalized cognitive decline among older adults. Importantly, the mCoDy score contains much more information than the binary diagnosis of MCI (scored as yes or no) and allows a quantitative statistical approach. In addition, this score shows that some older adults suffer from objective dysfunctions in episodic memory, and not only from subjective feelings of cognitive impairment.

The above findings extend previous papers suggesting that MCI is often characterized by a significant impairment in episodic memory, which is the ability to recall specific events, situations, and experiences ^59^. Even though episodic memory impairment is a characteristic of aMCI, not everyone with episodic memory problems will develop aMCI ^59^. Individuals with MCI often exhibit deficits in verbal fluency tasks ^7^, which is the cognitive function involving the ability to retrieve and produce words quickly based on specific criteria, such as a particular letter or a category ^60^. Nevertheless, in our study, the VFT scores could not be combined with the MMSE, MoCa, WLM and WLR scores into one factor because the loading of VFT scores on the first factor were much too low. As expected, we found that the mCoDy score was only partially (19.0%) explained by increasing age and lower levels of education (see also introduction). This leaves a significant proportion of the variance unexplained.

Importantly, in the present study we were unable to detect any correlation between the quantitative DSOA and mCoDy scores, indicating that both dimensions are independently from each other. Consequently, the DSOA score represents a novel subclinical concept even in the absence of a major depressive episode, BPSD and ADL dysfunctions. Nevertheless, we observed that Petersen’s item 1 was correlated with the DSOA score. This suggests that the subjective perception of cognitive deterioration may be linked with heightened distress and affective scores in older adults ^61^. This may be explained by the theory that the subjective feeling of cognitive impairment is indeed a distress for older adults ^62–64^.

### Effects of ACEs on the DSOA and mCoDy

The third major finding of this study is that ACE and NLE (especially health and money) have a highly significant impact on the DSOA, and no effects at all on the mCoDy domain scores. ACE and NLE are significant risk factors for depression and other psychological disorders in adulthood ^23,24,65^. Kraaij, Arensman and Spinhoven ^27^ reported that almost all NLE have a small but significant association with depression. The total number of negative life events and everyday problems appeared to have the strongest link with depression ^27,66^. Consequently, our results indicate that the DSOA dimensional score reflects in part the distress that originates from relevant NLE and that previous ACE may cause together with NLE a much stronger impact on the DSOA score. As such, the combination of ACE, NLE and subjective feelings of cognitive impairments are together major predictors of the increasing DSOA scores in older adults. This perspective acknowledges the impact of negative life experiences (ACE and NLE) on mental health and proposes that DSOA scores (comprising the neuroticism trait) may be partly determined by the individual’s resilience and coping mechanisms.

The final question is then whether DSOA should be regarded either as a psychopathological dimension or as a human response to environmental stressors, such as NLE and negative formative experiences. The first viewpoint considers that DSOA are not a normal part of aging, but rather a sign of pathology that requires treatment or clinical intervention. On the other hand, if these symptoms are seen as a response to environmental stressors only, it implies that they are a normal reaction to the challenges and hardships that an individual has encountered throughout their life. Clinicians could use the subjective DSOA complaints as a criterium whether psychotherapeutic intervention is indicated or not. Alternatively, future biomarker research should determine whether the DSOA have pathway substrates indicative of physiological stress (e.g., cortisol axis, immune activation, oxidative stress) that should be targeted by novel treatments.

Regarding the mCoDy score, prior research findings may require revision, because previous suggestions that NLE may accelerate cognitive decline in older adults ^12,13^ may not be correct after excluding subjects with clinical depression (this study).

Also, studies showing that ACE are substantially correlated with cognitive impairments in older adults ^9–11^ cannot be confirmed after excluding subjects with major depression (this study). In this respect, one study reported that ACE had a negative effect on cognitive function in middle-aged and older adults and that depression mediated this relationship ^23,67^. However, since we excluded individuals with major depression, no such mediating effects may be observed.

### DSOA and mCoDy clusters

This study’s fourth significant finding is that cluster analysis identified three clusters of people based on mCoDy and DSOA scores and that these classes do not align with the clinically defined distinction between aMCI and healthy controls. Thus, three cases diagnosed with aMCI were allocated to the healthy control group, while three healthy controls were assigned to the aMCI group. In addition, the cluster analysis uncovered a new cluster that was shaped by DSOA rating scales, and part of the aMCI cases were allocated to that new cluster. The DSOA cluster included healthy controls (70.96%) and individuals with aMCI (29.04%). As such, cluster analysis derived two classes of individuals, a first with DSOA, and another with mCoDy. The first cluster showed much higher scores on all DSOA rating scales (except TGDS), and the latter cluster showed lower cognitive scores on MMSE, MoCA, WLM, WLR and VFT.

Without computing the DSOA scores and without cluster analysis we would never have been able to uncover that part of aMCI individuals and “healthy controls” may be allocated to a DSOA class. Our results indicate that the clinical diagnosis based on Petersen criteria is overinclusive, as it includes subjects who belong to the DSOA group. The latter cases might be clinical misclassifications, whereby cases with DSOA may mistakenly be classified as aMCI, presumably because they also exhibit subjective cognitive symptoms (Petersen item 1), although neuropsychological testing did not show any objective signs of cognitive decline. In addition, previous papers have demonstrated that the Petersen criteria are overinclusive ^7,8^. As a result, we propose the term “mild cognitive dysfunction” (mCoDy) as an alternative to the overinclusive aMCI diagnosis, and the quantitative mCoDy score as a better alternative than the incorrect (as overinclusive) binary (limited information) aMCI diagnosis.

All in all, this study clarified why the diagnosis of aMCI according to the Petersen criteria is overinclusive, because this diagnosis includes subjects with DSOA who have subjective feelings of cognitive difficulties but lack objective signs of cognitive decline.

## Limitations and future research

The current results deserve replication in individuals without clinical depression and ADL dysfunctions in other countries and cultures. Future research should examine whether the DSOA and mCoDy scores can be externally validated by any physiological stress-associated pathways that may impact affect or neurocognition, including immune, oxidative stress, neurotrophic, and neuroplasticity pathways. In addition, longitudinal studies are needed to explore long-term outcomes of DSOA.

## Conclusion

The study concludes that in older adults without major depression and ADL dysfunctions, mCoDy and DSOA are two distinct dimensions. The DSOA dimensions is influenced by ACE, NLE, and the subjective feeling of cognitive decline, whereas the mCoDy score is affected by aging and lower education. This distinction is crucial for understanding and addressing mental health issues in older adults without major depression. The results also highlight the long-term effects of ACE in early life and NLE on mental health in older adults. The clinical impact of the current study is that clinicians should carefully screen older adults for DSOA after excluding a major depressive episode and BPSD. This may be important for the treatment of DSOA versus mCoDy. While the former is associated with psychological trauma and psychosocial stressors, psychotherapeutic interventions may be needed, whereas mCoDy might be treatable instead with lifestyle interventions, including physical activity ^68^ and perhaps specific antioxidant treatments targeting the oxidative stress pathways ^69^.

## Availability of data and materials

The dataset generated during and/or analysed during the current study will be available from MM upon reasonable request and once the authors have fully exploited the dataset.

## Author’s contributions

All authors contributed to the paper. MM, CT, and V-LT-C: conceptualization and study design. V-LT-C and MM: first draft writing. V-LT-C, MM, CT, SH, GN, and MS: editing. GN, CT and V-LT-C: recruitment of patients. MM and V-LT-C: Statistical analyses. All authors revised and approved the final draft.

## Institutional Review Board Statement

This study was approved by the Institutional Review Board (IRB) of the Faculty of Medicine, Chulalongkorn University, Bangkok, Thailand (IRB no. 0372/65), which complies with the International Guideline for Human Research Protection as required by the Declaration of Helsinki.

## Informed consent

Before taking part in the study, all participants and/or their caregivers provided written informed consent.

